# Potential risk polarization for acute myocardial infarction during the COVID-19 pandemic: Single-center experiences in Osaka, Japan

**DOI:** 10.1101/2022.10.28.22281657

**Authors:** Masato Furui, Kenji Kawajiri, Takeshi Yoshida, Bunpachi Kakii, Norikazu Oshiro, Mai Asanuma, Hiroaki Nishioka, Hideichi Wada

## Abstract

This study compared the time course and outcomes of acute myocardial infarction, including mechanical complications and hospital mortality, before and after the coronavirus disease 2019 (COVID-19) pandemic at a regional core hospital in South Osaka, Japan. Moreover, it identified predictors for hospital mortality and mechanical complications. In total, 503 patients who underwent emergency percutaneous coronary intervention between January 2011 and December 2021 at our institution were examined retrospectively. The time course of acute myocardial infarction, mechanical complications, and mortality rate before and after the COVID-19 emergency declaration were compared. Overall, 426 patients with ST-segment elevation myocardial infarction and 77 patients with non-ST-segment elevation myocardial infarction were identified. For patients with ST-segment elevation myocardial infarction, the onset-to-door time was longer (181 vs. 156 min, P = 0.001) and mechanical complications were worse (7.8% vs. 2.6%, P = 0.025) after the emergency declaration of COVID-19 than before the pandemic. Age, low ejection fraction, out-of-hospital cardiac arrest, and mechanical complications were identified as independent risk factors for hospital mortality among patients with ST-segment elevation myocardial infarction, using multivariable analysis. Post-declaration, age, walk-ins, referrals, and intra-aortic balloon pump use were independent predictors of mechanical complications among patients with ST-segment elevation myocardial infarction. Onset-to-door time and mechanical complication rate increased after the COVID-19 declaration among patients with ST-segment elevation myocardial infarction. Arrival by walk-in and a referral that caused treatment delay were identified as independent risk factors for mechanical complication, in addition to age, use of intra-aortic balloon pump, and post-declaration of COVID-19. Therefore, the risks posed by the COVID-19 pandemic might have a polarization tendency resulting from the relief or worsening of cardiac symptoms.

## Introduction

The coronavirus disease 2019 (COVID-19) pandemic has been in existence since 2020. The infection situation is not much different in Japan, and the nation has already experienced the sixth wave of COVID-19. Common behavioral patterns have been adjusted to reduce infection risk, and patients are being asked to avoid population-dense areas for the same reason. Accordingly, a reduction in hospitalizations for acute myocardial infarction (AMI) has been reported. [1–6] Although the impact of the COVID-19 pandemic on door-to-balloon time and mechanical complications of AMI have been previously examined in other countries, such reports are still scarce in Japan. [7–10]

A few studies on percutaneous coronary intervention (PCI) impact in Tokyo, Japan, have been reported; however, whether these are relevant to other regions is unknown due to the rural-urban emergency care disparity. [11,12] For example, South Osaka does not include a metropolitan area, being the second largest prefecture in Japan; therefore, accessibility to PCI-capable institutions will be different from that in Tokyo. [13,14] After the pandemic, studies considering consultation forms, such as walk-ins, direct arrival by emergency medical service (EMS), or referral transport from other non-PCI-capable facilities, are rare. [14]

Therefore, this study compared the time course and outcomes of AMI, including mechanical complications and in-hospital mortality, before and after the COVID-19 pandemic declaration at a regional core hospital in South Osaka, Japan. Moreover, this study examined whether differences in types of hospital visits were a predictor of the outcomes.

## Materials and Methods

### Study design and population

The study protocol conformed to the ethical guidelines of the Declaration of Helsinki and was approved by the Ethics Committee of the Matsubara Tokushukai Hospital (approval number: 22-01). The informed consent requirement from patients was waived due to the retrospective nature of this study. The corresponding author (Masato Furui) has full access to all data in this study and takes responsibility for data integrity and analysis.

This study was a retrospective observational cohort study conducted on patients who underwent emergency PCI for AMI at the Matsubara Tokushukai Hospital in Osaka, Japan, between January 2011 and December 2021. The medical records of 796 patients with acute coronary syndrome (ACS) were reviewed, and data of 503 AMI patients were extracted after excluding those with recent myocardial infarction (MI) with an onset-to-door time >14 days and unstable angina pectoris. The participants were classified into ST-segment elevation myocardial infarction (STEMI) and non-STEMI (NSTEMI) groups. The Japanese government declared a state of emergency on April 7, 2020, following the World Health Organization’s initial declaration of COVID-19 as a pandemic on March 11, 2020. [15,16] With this emergency declaration as a borderline, the characteristics, time course, and outcomes were compared before and after the declaration to evaluate the impact of the COVID-19 pandemic on both the STEMI and NSTEMI groups (Figure 1).

**Fig 1.**
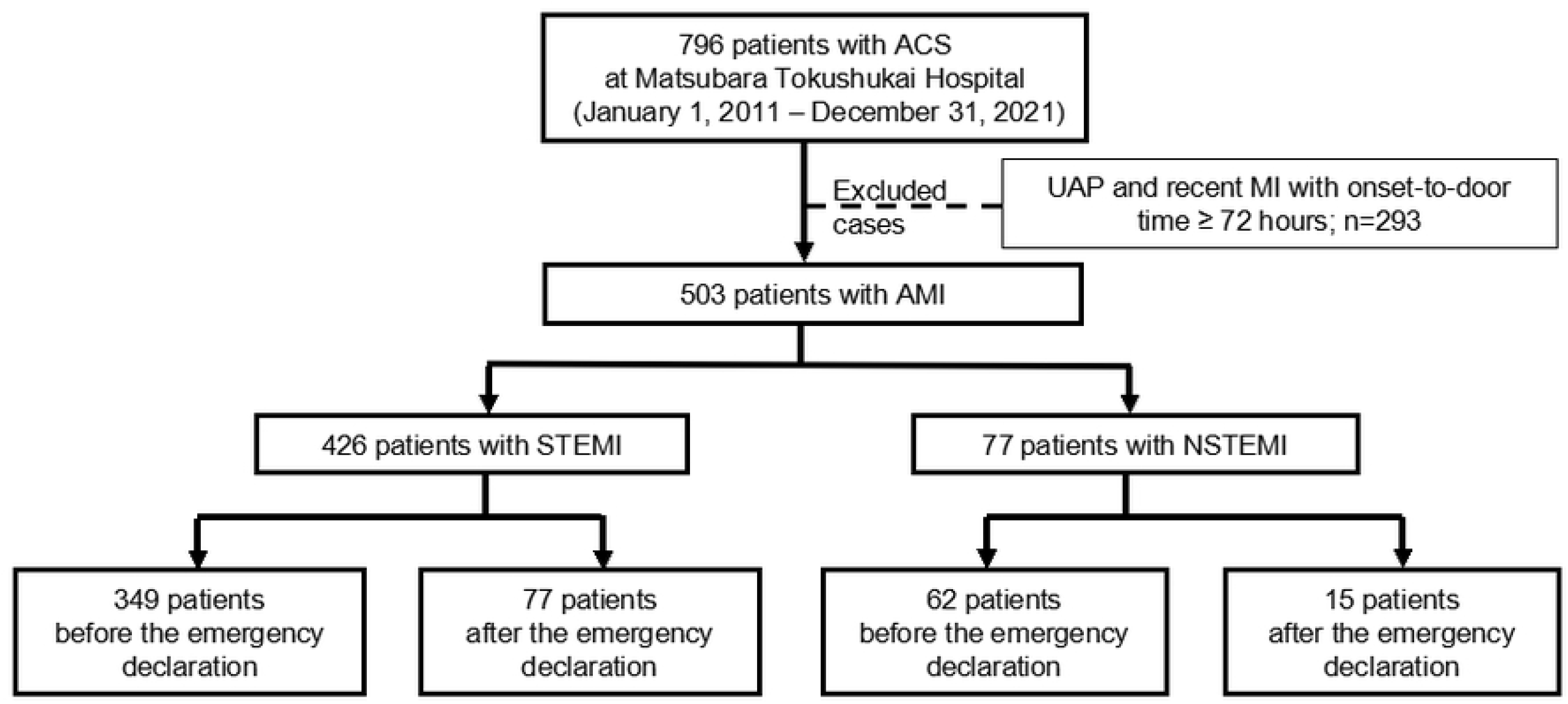
Flow chart illustrating the selection of the study participants. Abbreviations: ACS, acute coronary syndrome; AMI, acute myocardial infarction; MI, myocardial infarction; NSTEMI, non-ST-segment elevation myocardial infarction; STEMI, ST-segment elevation myocardial infarction; UAP, unstable angina pectoris.

### Definitions

AMI was defined based on the fourth universal definition of MI, and ACS was defined according to the Japanese Circulation Society guidelines. [17,18] Infarction was classified into STEMI or NSTEMI, depending on the presence or absence of ST-segment elevation. Onset-to-door time and door-to-balloon time were defined as the time from symptom onset to hospital arrival and from hospital arrival to balloon dilation or thrombus aspiration, respectively. [19] In the present study, renal dysfunction was defined as creatinine level >1.5 mg/dL. Left ventricular ejection fraction (LVEF) was assessed using echocardiography. Primary PCI was defined as urgent balloon angioplasty (with or without stenting) without employing fibrinolytic therapy to open the infarct-related artery.

The primary outcomes were onset-to-balloon time and in-hospital mortality. The secondary outcome was the incidence of mechanical complications. Therefore, we compared onset-to-door time, mechanical complications, and hospital mortality between patients before and after the COVID-19 emergency declaration in Japan. Mechanical complications, including cardiac rupture, ventricular septal perforation, and papillary muscle rupture, were assessed using echocardiography. [20–22]

### Statistical analysis

Categorical variables were expressed as numbers and proportions, whereas continuous variables were presented as mean±standard deviation. Student’s t-test, chi-squared test, and Mann–Whitney U test were utilized for other statistical analyses. Multivariate logistic regression analysis was performed for risk factors associated with hospital mortality and mechanical complications in patients with STEMI. This analysis considered age, walk-ins, referrals, out-of-hospital cardiac arrest (OHCA), LVEF, single-vessel disease, anterior MI as a culprit, onset-to-door time, intra-aortic balloon pump (IABP) use, mechanical complications, and post-COVID-19 declaration, as possible predictors. [7,22,23] The odds ratios (OR) and 95% confidence intervals (CI) were calculated. The number of patients who underwent venoarterial extracorporeal membrane oxygenation was few; therefore, it was not entered because we could not effectively analyze it. Moreover, NSTEMI analysis was not valid due to the small number. All statistical analyses were performed using JMP^®^ version 9.0 (SAS Institute Inc., Cary, NC), and differences were considered statistically significant at a *P*-value <0.05.

## Results

We extracted the data of 796 patients who required emergency PCI for ACS from medical records. Of these, 503 AMI patients were identified as participants in this study (Figure 1), of whom 426 and 77 patients were categorized into the STEMI and NSTEMI groups, respectively. Table 1 summarizes the basic characteristics, including the patients’ background and pre-PCI examination findings before and after the COVID-19 declaration in each group. In the STEMI group, the number of patients with Killip classes III–IV was significantly higher after the declaration than it was before the declaration (12.0% [42/349] vs. 21.3% [16/77], *P*=0.010). Although the prevalence of OHCA was not significantly different, it tended to be higher after the declaration (2.3% [8/349] vs. 6.5% [5/77], *P*=0.052). There were no significant differences in comorbidities, clinical presentation, and laboratory data before and after the declaration, except for Killip III–IV classification in the STEMI group, as previously described.

**Table 1.**
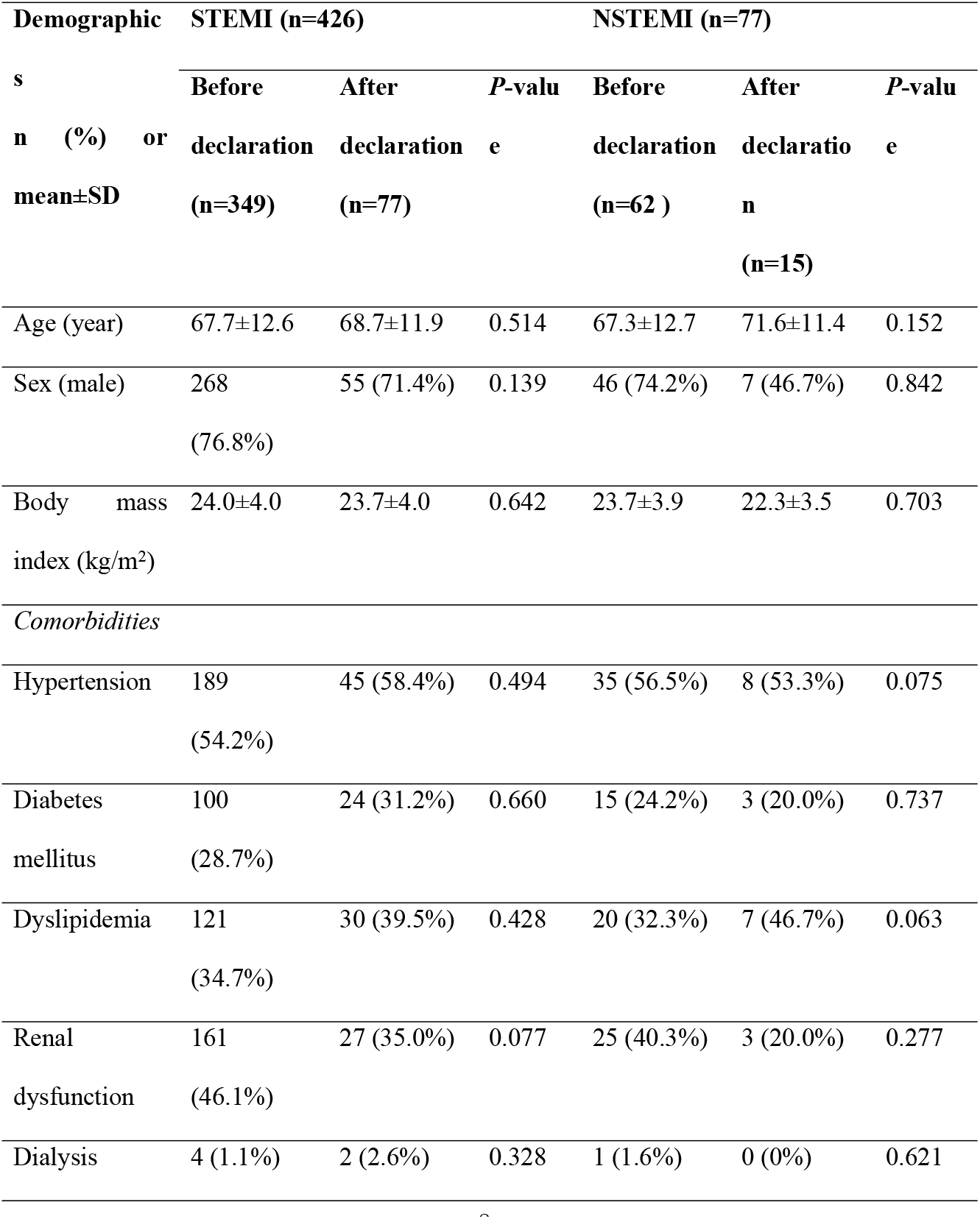
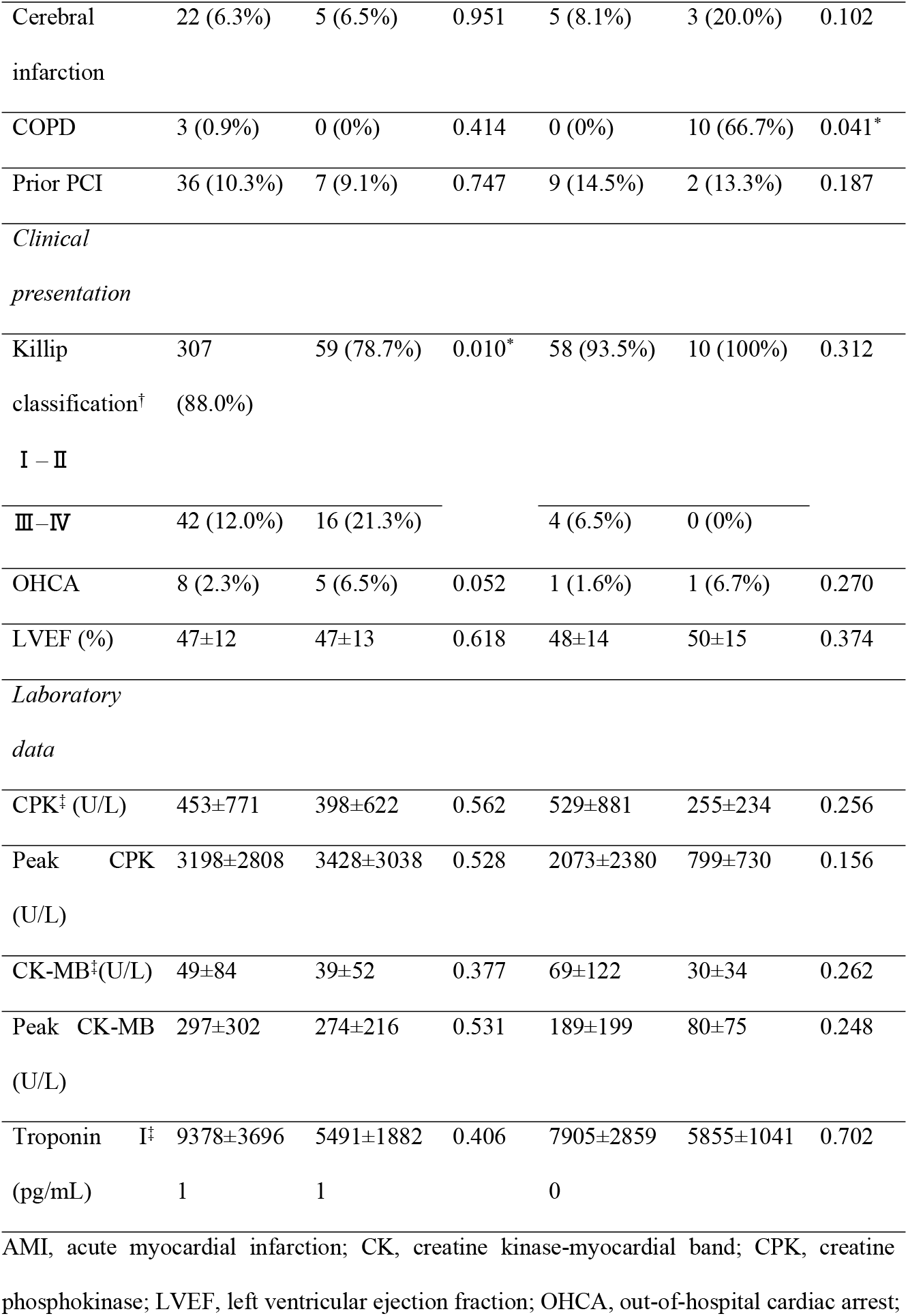
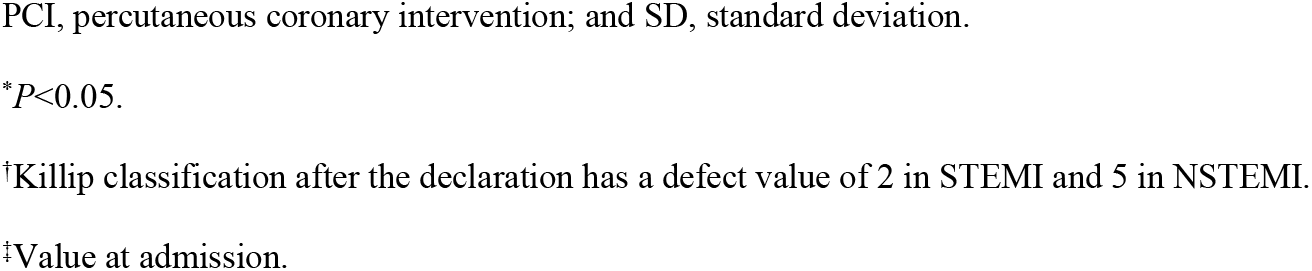
Basic characteristics of AMI patients before and after the COVID-19 declaration.

Table 2 shows consultation types, (such as walk-ins, arrival by EMS, and referrals,) the time course, and outcomes before and after the declaration in each group. In the STEMI group, there were fewer referred patients after the declaration than there were before the declaration. However, there was no significant difference in the door-to-balloon time before and after the declaration (109±129 vs. 96±54 min, *P*=0.420). Onset-to-door time (a primary outcome) in patients with STEMI was significantly longer after the COVID-19 declaration than it was before the declaration (156±134 vs. 181±145 min, *P*=0.001).

**Table 2.**
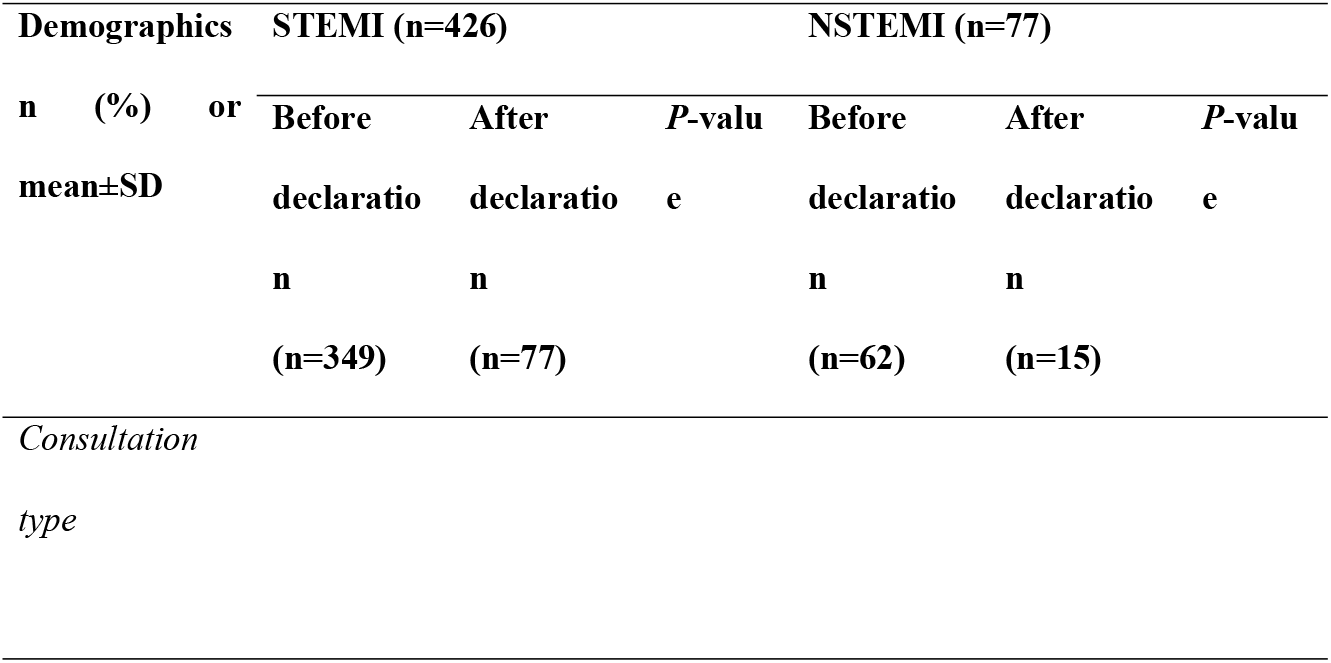
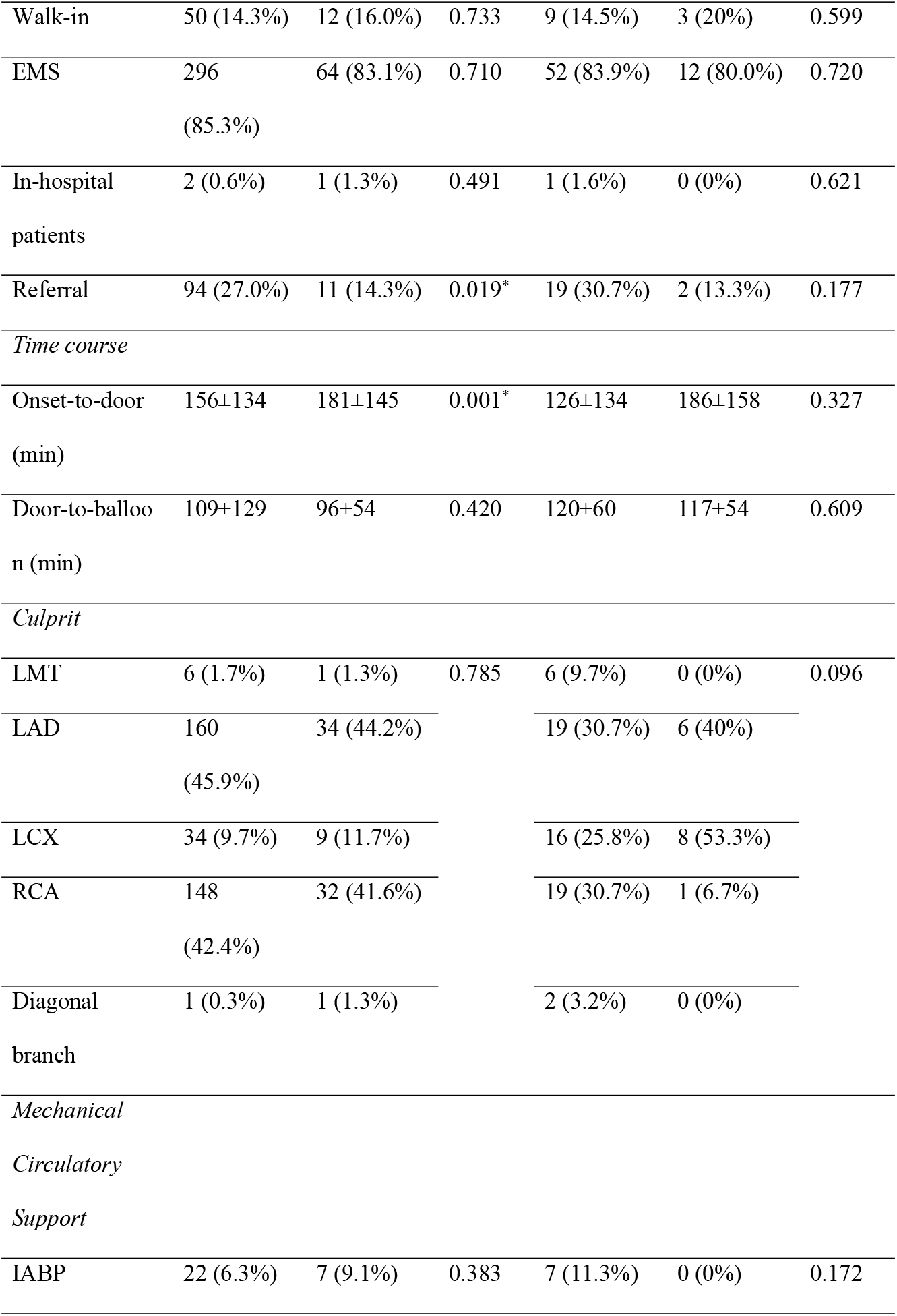
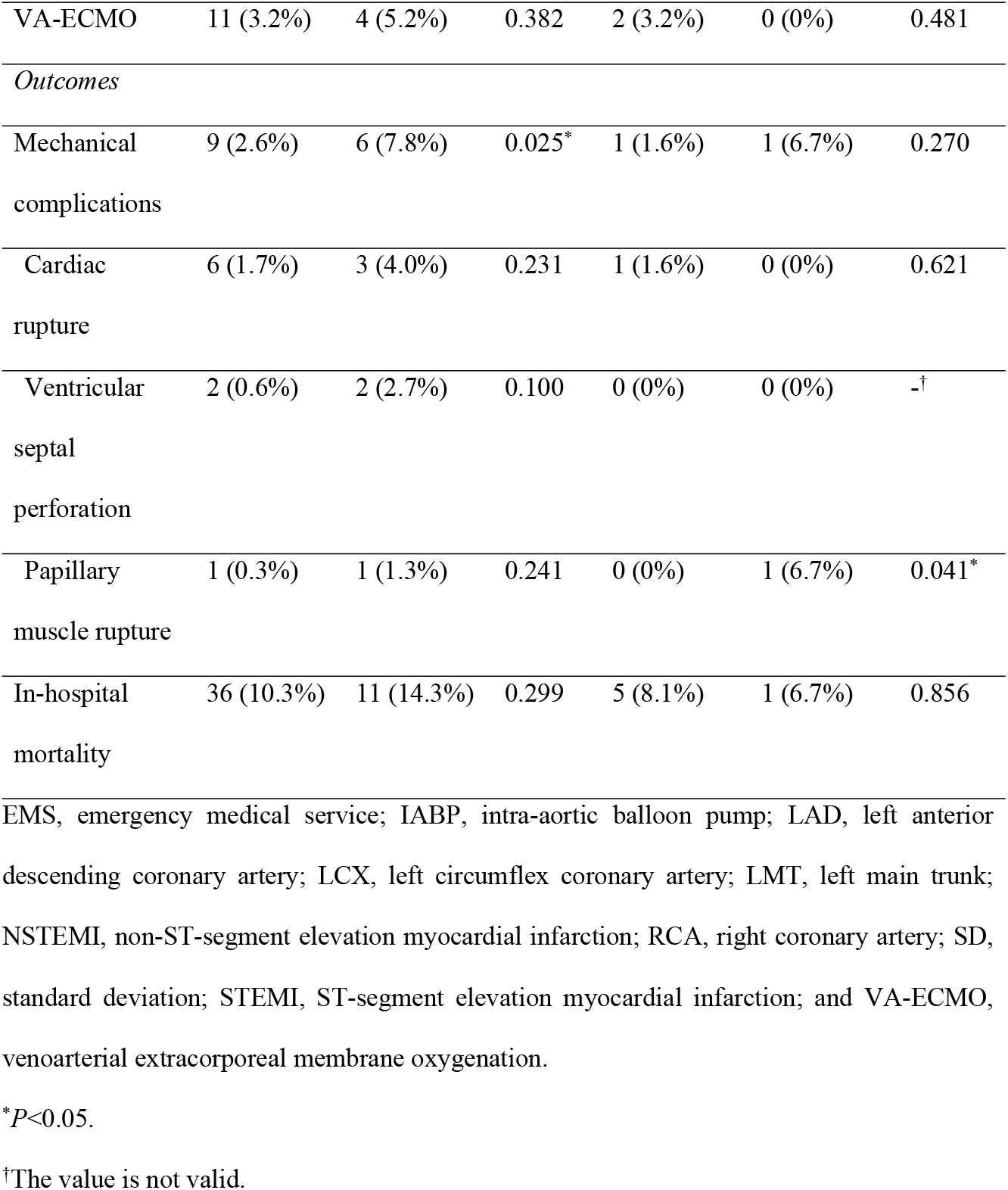
Consultation type, time course, and outcomes in AMI patients before and after the declaration.

Hospital mortality, another primary outcome, occurred in 11.0% (47/42 6) of patients with STEMI and 7.8% (6/77) of patients with NSTEMI. Contrarily, mechanical complications, a secondary outcome, occurred in 3.5% (15/426) of patients with STEMI patients and 2.6% (2/77) of patients with NSTEMI. The mechanical complication rate was significantly higher after the COVID-19 declaration than it was before the declaration (2.6% [9/349] vs. 7.8% [6/77], *P*=0.025) in the STEMI group; however, there was no significant difference in hospital mortality (10.3% [36/349] vs. 14.3% [11/77], *P*=0.299).

Table 3 presents the results of the multivariate analysis for the primary outcomes. Age (1-year increase: OR 1.087, 95% CI 1.045–1.135, *P* ≤ 0.0001), OHCA (OR 61.883, 95% CI 9.726–595.506, *P*<0.0001), LVEF (1% increase: OR 0.943, 95% CI 0.911–0.975, *P*=0.0004), IABP use (OR 10.523, 95% CI 3.544–33.544, *P* ≤ 0.0001), and mechanical complications (OR 10.724, 95% CI 2.233–57.236, *P*=0.003) were established as independent predictors of hospital mortality. Table 4 presents the results of the multivariate analysis for the secondary outcome. Age (1-year increase: OR 1.115, 95% CI 1.037–1.221, *P*=0.0019), walk-in visits (OR 14.695, 95% CI 3.265–80.468, *P*=0.0005), referral visits (OR 4.854, 95% CI 1.050–25.003, *P*=0.043), IABP use (OR 29.094, 95% CI 4.209–255.552, *P*=0.0008), and post-COVID-19 declaration (OR 5.006, 95% CI 1.131–22.443, *P*=0.035) were identified as independent predictors of mechanical complications.

**Table 3.**
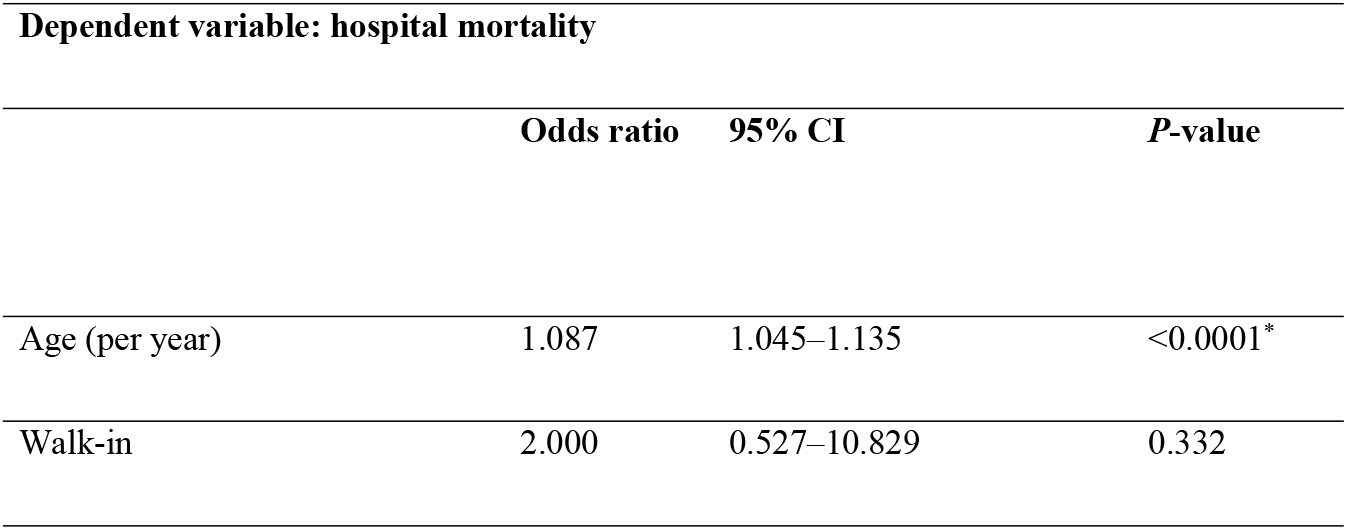
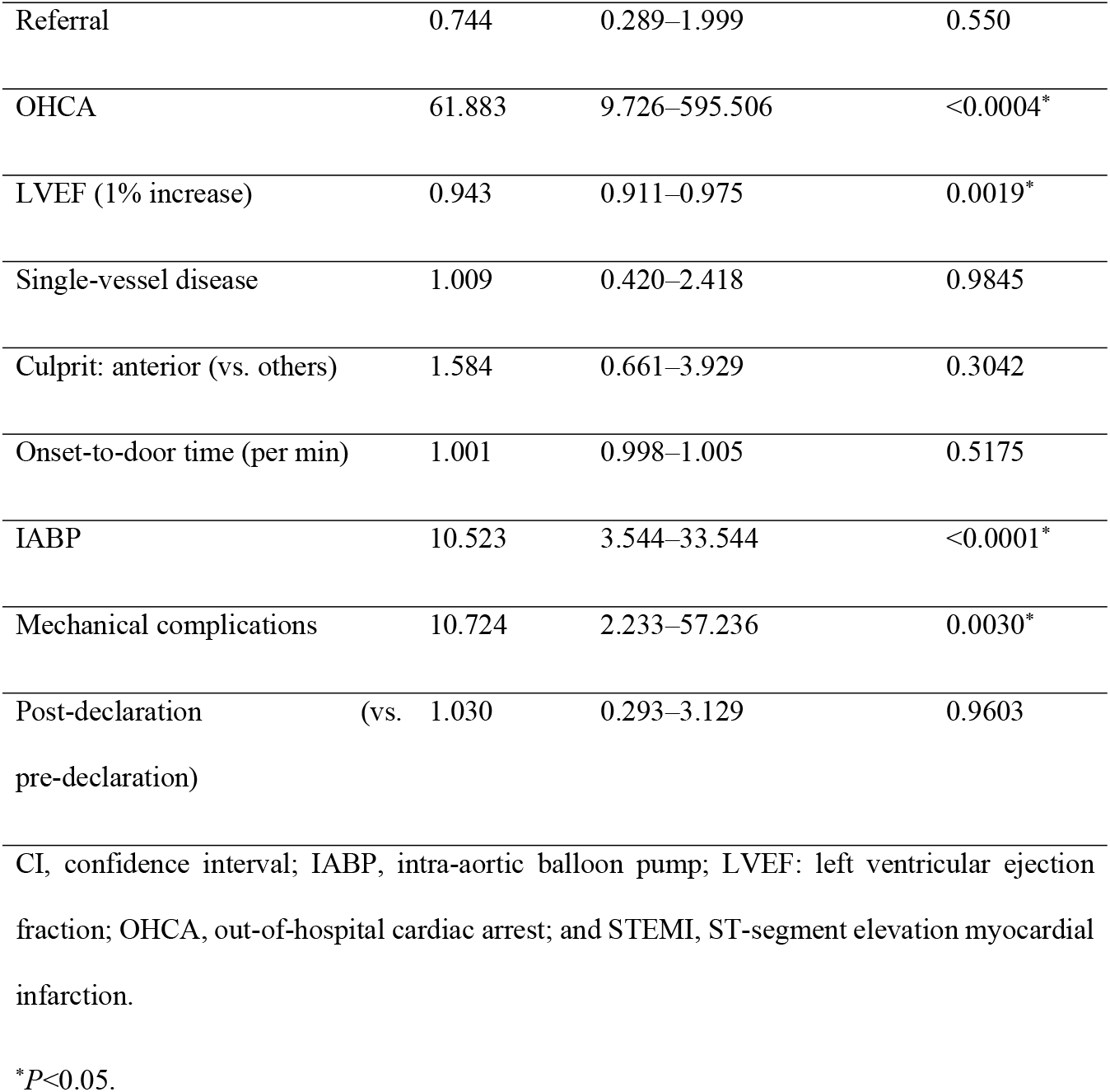
Multivariate logistic regression analysis of risk factors for STEMI. Dependent variable: hospital mortality.

| **Dependent variable: hospital mortality** |  |  |  |
| --- | --- | --- | --- |
|  | **Odds ratio** | **95% CI** | ***P*-value** |
| Age (per year) | 1.087 | 1.045–1.135 | <0.0001* |
| Walk-in | 2.000 | 0.527–10.829 | 0.332 |
| Referral | 0.744 | 0.289–1.999 | 0.550 |
| OHCA | 61.883 | 9.726–595.506 | <0.0004* |
| LVEF (1% increase) | 0.943 | 0.911–0.975 | 0.0019* |
| Single-vessel disease | 1.009 | 0.420–2.418 | 0.9845 |
| Culprit: anterior (vs. others) | 1.584 | 0.661–3.929 | 0.3042 |
| Onset-to-door time (per min) | 1.001 | 0.998–1.005 | 0.5175 |
| IABP | 10.523 | 3.544–33.544 | <0.0001* |
| Mechanical complications | 10.724 | 2.233–57.236 | 0.0030* |
| Post-declaration<br>(vs. pre-declaration) | 1.030 | 0.293–3.129 | 0.9603 |

CI, confidence interval; IABP, intra-aortic balloon pump; LVEF: left ventricular ejection fraction; OHCA, out-of-hospital cardiac arrest; and STEMI, ST-segment elevation myocardial infarction.
\* $P < 0.05$ .

**Table 4.**
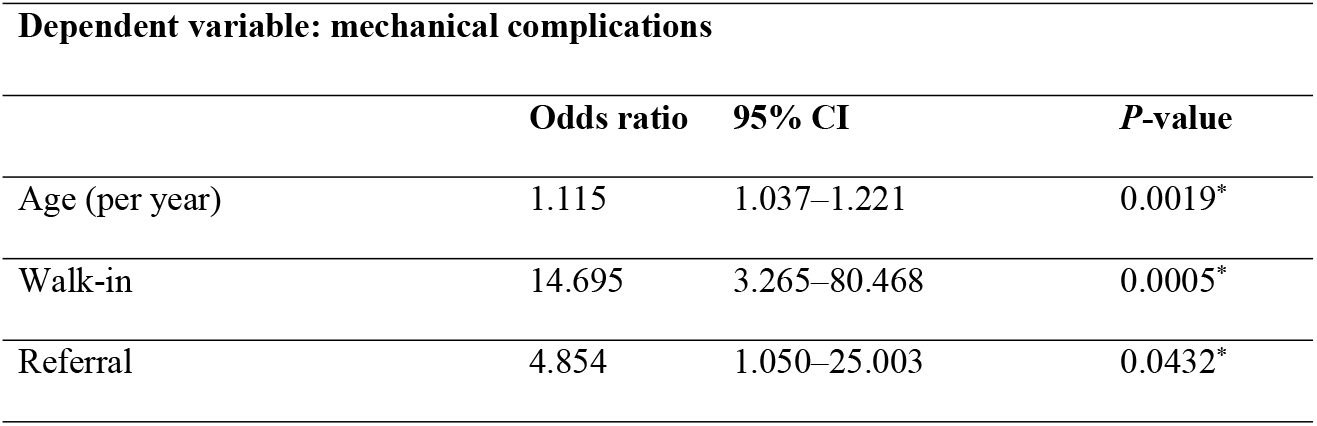
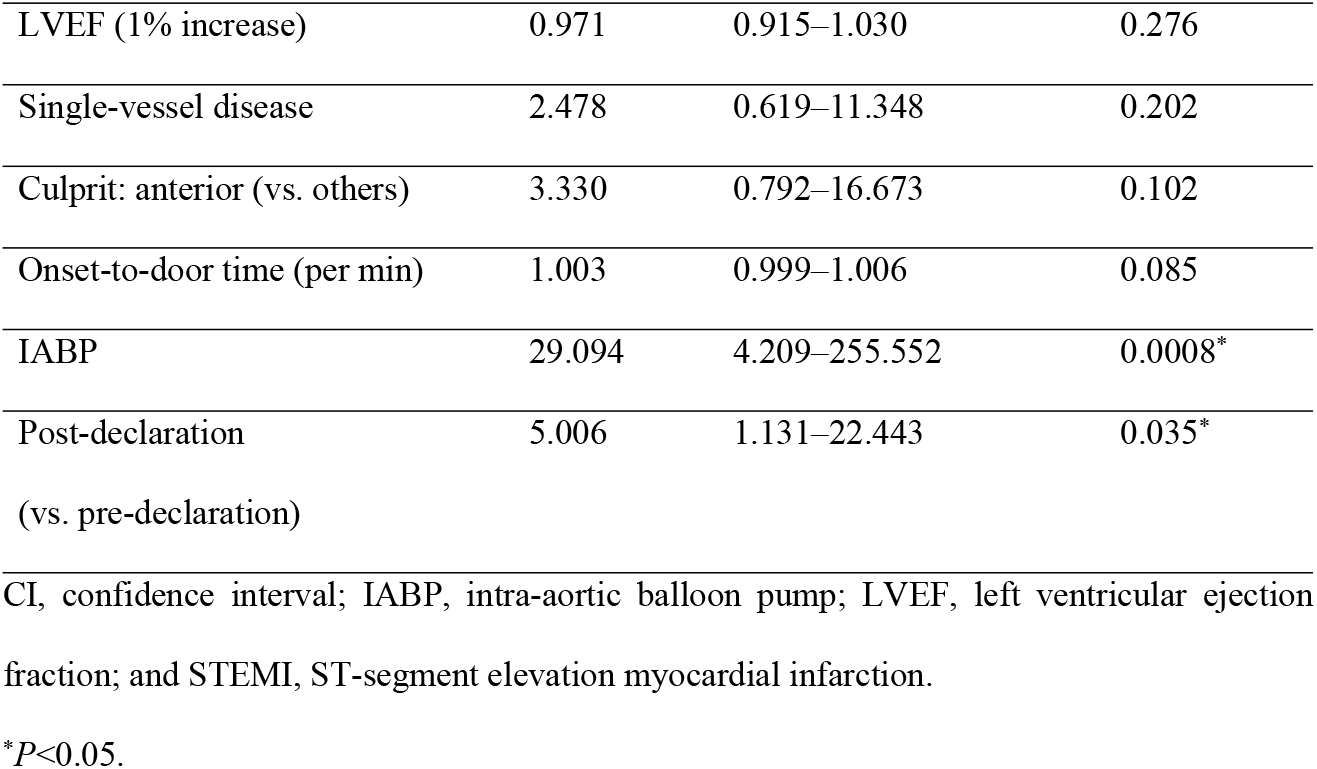
Multivariate logistic regression analysis of risk factors for STEMI. Dependent variable: mechanical complications.

| Dependent variable: mechanical complications |  |  |  |
| --- | --- | --- | --- |
|  | Odds ratio | 95% CI | *P*-value |
| Age (per year) | 1.115 | 1.037–1.221 | 0.0019* |
| Walk-in | 14.695 | 3.265–80.468 | 0.0005* |
| Referral | 4.854 | 1.050–25.003 | 0.0432* |
| LVEF (1% increase) | 0.971 | 0.915–1.030 | 0.276 |
| Single-vessel disease | 2.478 | 0.619–11.348 | 0.202 |
| Culprit: anterior (vs. others) | 3.330 | 0.792–16.673 | 0.102 |
| Onset-to-door time (per min) | 1.003 | 0.999–1.006 | 0.085 |
| IABP | 29.094 | 4.209–255.552 | 0.0008* |
| Post-declaration<br>(vs. pre-declaration) | 5.006 | 1.131–22.443 | 0.035* |

CI, confidence interval; IABP, intra-aortic balloon pump; LVEF, left ventricular ejection fraction; and STEMI, ST-segment elevation myocardial infarction.
\* $P < 0.05$ .

Table 5 shows the association of walk-ins and referrals with the time course after the declaration in patients with STEMI. Regarding arrival by walk-in, both onset-to-door and door-to-balloon times were significantly extended in patients with walk-in arrivals, compared with those from non-walk-in arrivals (175 vs. 127 min, *P*=0.015 and 150 vs. 95 min, *P*=0.038). Conversely, referral patients had a significantly longer onset-to-door time (164 vs. 124 min, *P*=0.016) and shorter door-to-balloon time than non-referral patients (87 vs. 113 min, *P*=0.009).

**Table 5.**
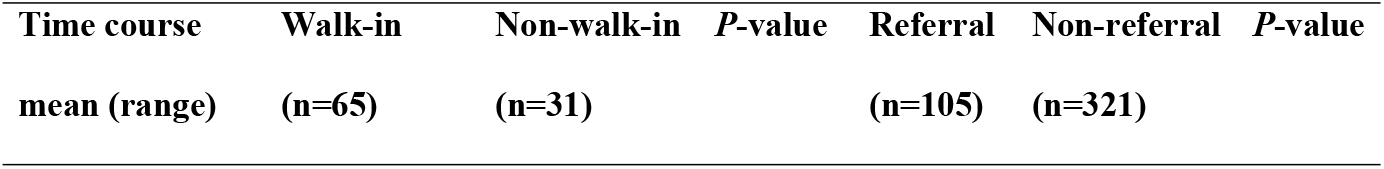
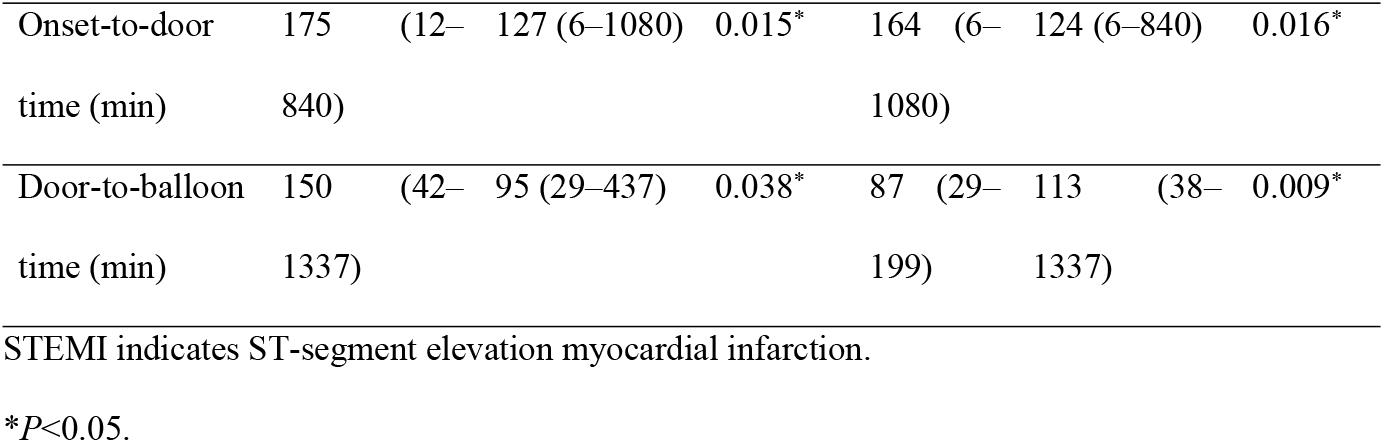
The association of walk-ins and referrals with time course after COVID-19 declaration in the STEMI group.

## Discussion

Several studies have reported the impact of the COVID-19 pandemic on AMI in Tokyo. [3,7] However, our hospital’s secondary medical service area in South Osaka is distinct from that in Tokyo and has an unevenly distributed population, similar to hospitals, especially PCI-capable facilities. [13,24,25] Moreover, arrival by walk-in or referral as risk factors for AMI have rarely been discussed following the COVID-19 pandemic. Therefore, our study investigated the influence of walk-ins and referrals on mechanical complications and hospital mortality to address this and compare time course and outcomes before and after the COVID-19 pandemic in STEMI and NSTEMI groups. We established that onset-to-door time was prolonged and mechanical complications developed more frequently in STEMI patients after the COVID-19 declaration, with no significant difference between both groups. These findings suggest that, in addition to post-COVID-19 declaration, walk-ins and referrals were risk factors for mechanical complications.

Regarding risk factors for hospital mortality, it was reported that patients with lower LVEF have higher mortality rates and develop heart failure easily. [26,27] Once mechanical complications occur, they can fatally affect patients’ hemodynamics. Although various operative procedures and strategies in surgery timing have been devised, operative mortality remains high. [21,22,28] Therefore, investigating the risk factors and preventing subsequent complications may be important in reducing associated mortality.

Several risk factors for mechanical complications are known; previous studies have shown that established mechanical complication risk factors include age, female sex, anterior MI, de novo MI, and single-vessel disease. However, the analysis conducted in this study did not identify single-vessel disease and anterior MI as mechanical complication risk factors. [21,22] Moreover, once mechanical complications occur, patients often experience cardiogenic shock, thus requiring IABP use, which may naturally become a risk factor.

Considering previous findings, the identification of walk-ins and referrals as risk factors for mechanical complications are unique points in the present study. Walk-in patients often have mild symptoms, making this seem contradictory. However, in some cases with mild symptom presentation, patients’ hearts can suffer from unnoticed load until AMI diagnosis, even after visiting the hospital because of unrestricted patient mobility or activity. Failure to limit this activity may become a mechanical complication risk. Thus, the time course for walk-in patients is distinct from that for patients transported by ambulance. Patients who notify the EMS are likely to immediately undergo consultation and check-ups by emergency doctors who begin monitoring as soon as the patient arrives at the hospital. Useful information by rescue crews generally aids in smooth AMI diagnosis. Conversely, walk-in patients have to wait longer for consultation or check-ups. Therefore, there may be a pronounced time lag from their arrival to diagnosis or PCI, compared with that of patients who notify EMS.

Referred patients were reported to have a prolonged onset-to-door time because they came from a nearby clinic or were transferred from non-PCI-capable facilities. [14,19] Previous studies reported that the time interval from an AMI onset to ventricular septal perforation manifestation had a bimodal distribution; therefore, a longer ischemic time can become a risk factor for mechanical complications due to interactions with unknown factors. [10,22] In contrast, there was a significantly shorter door-to-balloon time in referred patients with STEMI after the pandemic declaration. Although we could not specify the reason, preparing for PCI in advance by referral request and an improved acceptance system by the regional cooperation office might achieve a shorter door-to-balloon time. In any case, because a referral is indispensable due to the surrounding non-PCI-capable facilities, we suggest that an effective EMS system and crew education on carrying suspected patients to PCI-capable facilities is important. [11,13,14]

In the STEMI group, OHCA rates tended to be higher after the declaration than it was before, although this failed to reach statistical significance. Given the potential for a higher OHCA rate and risk factors for mechanical complications, such as walk-ins and referrals, AMI risk polarization may have emerged on the clinical scene during the COVID-19 pandemic. Although rare, spontaneous reperfusion occurs in 7–30% of patients with STEMI, as stated in previous studies. [29,30] In addition, patients who follow stay-at-home orders and initially tolerate or adapt to their symptoms may likely experience spontaneous reperfusion. Walk-in patients with apparently mild symptoms or referred patients with STEMI may be at risk of mechanical complications, which were identified in the present study as mortality risk in STEMI after the COVID-19 pandemic. Therefore, care should be taken regarding walk-in or referral patients suspected of STEMI.

Conversely, more severe symptoms can result in patients notifying EMS; however, their situation can become critical due to delays that may lead to OHCA during transport to PCI-capable facilities. The fear of contracting COVID-19 may have led to patient hesitation to visit hospitals, resulting in risk polarization (Figure 2). Timely arrival to PCI-capable facilities without hesitation is necessary for patients at risk of AMI.

**Fig 2.**
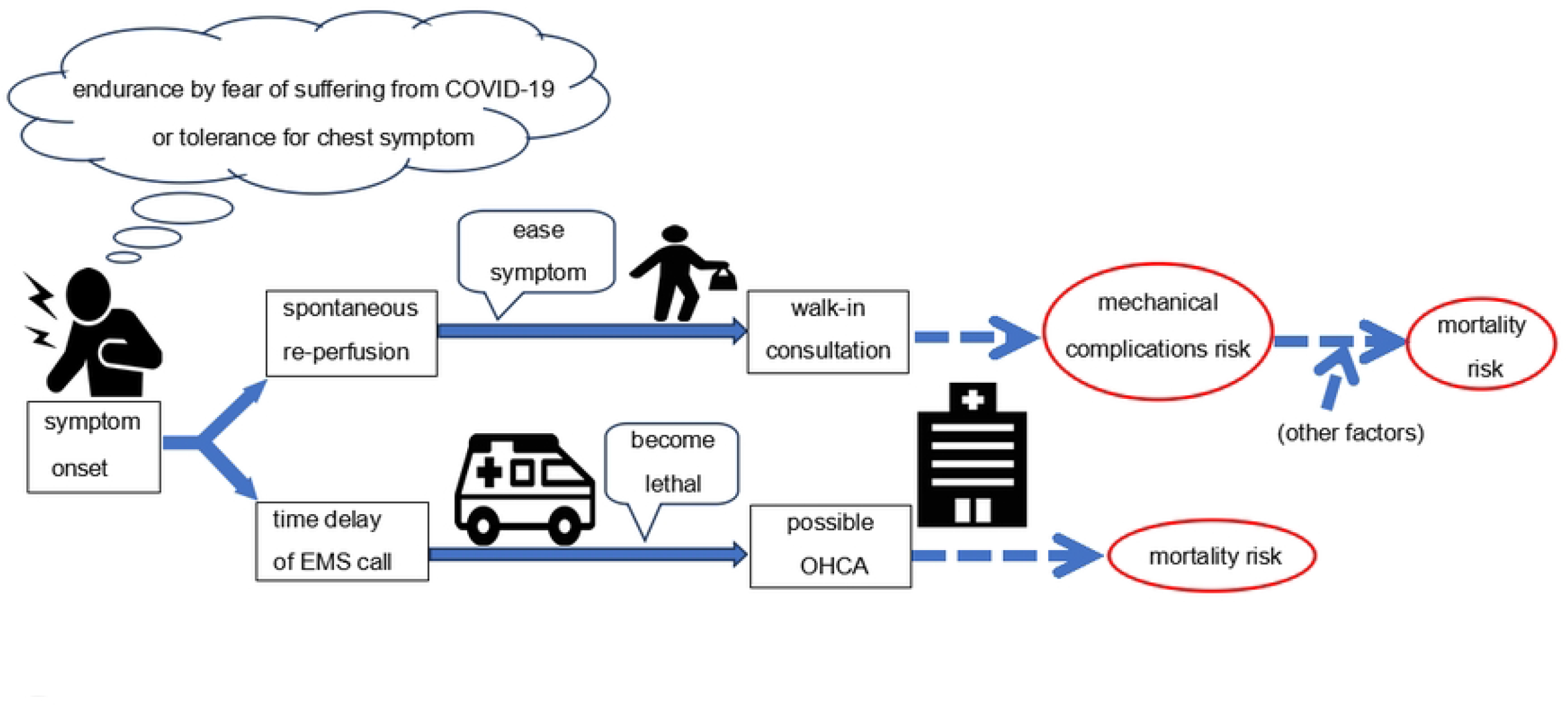
Polarization tendency of potential risks occurring from the patients’ psychology and possible time course after the COVID-19 pandemic. Abbreviations: EMS, emergency medical service; OHCA, out-of-hospital cardiac arrest.

The first limitation of this study is its retrospective observational design coupled with limited sample size from a single center. Our hospital covers only one of the areas in South Osaka. Thus, our data may not accurately represent Osaka as a whole because of emergency care or PCI volume disparity. [11–13] Second, additional data, encompassing EMS calls, transfer-to-door time, or COVID-19 examination waiting time, may be desirable in future studies to support the initial findings outlined in this study. Finally, the number of patients who died at home or during transport to our hospital was unknown and were excluded from the study population; therefore, OHCA and AMI prevalence rates may have been underestimated. Future research including these data will be required to further evaluate AMI-related mortality risk or mechanical complications during the COVID-19 pandemic.

## Conclusion

The onset-to-door time was prolonged and mechanical complications developed more frequently in patients with STEMI after the COVID-19 pandemic declaration than it did before. Age, low LVEF, OHCA, IABP use, and mechanical complications were established as independent risk factors for hospital mortality in patients with STEMI. Additionally, age, walk-ins, referrals, and IABP use were identified as independent predictors of mechanical complications in those with STEMI. Arrival by walk-in or referral, in patients with mild symptoms, influenced the outcomes of patients with AMI. Therefore, the risks posed by the COVID-19 pandemic might have a polarization tendency resulting from the relief or worsening of cardiac symptoms.

## Data Availability

All relevant data are within the manuscript and its Supporting Information files.

## Acknowledgments

We would like to thank Editage for English editing services.

## Notes

### Competing Interest Statement

The authors have declared no competing interest.

### Funding Statement

The authors received no specific funding for this work.

### Author Declarations

The Ethics Committee of the Matsubara Tokushukai Hospital (approval number: 22-01)

